# Low-density lipoprotein cholesterol levels are associated with poor clinical outcomes in COVID-19

**DOI:** 10.1101/2020.10.06.20207092

**Authors:** Álvaro Aparisi, Carolina Iglesias-Echeverría, Cristina Ybarra-Falcón, Iván Cusácovich, Aitor Uribarri, Mario García-Gómez, Raquel Ladrón, Raúl Fuertes, Jordi Candela, Williams Hinojosa, Carlos Dueñas, Roberto González, Leonor Nogales, Dolores Calvo, Manuel Carrasco-Moraleja, J. Alberto San Román, Ignacio J. Amat-Santos, David Andaluz-Ojeda

## Abstract

**Background:** Severe acute respiratory syndrome coronavirus 2 (SARS-CoV-2) is the sole causative agent of coronavirus infectious disease-19 (COVID-19).

**Methods:** We performed a retrospective single-center study of consecutively admitted patients between March 1^st^ and May 15^th,^ 2020, with a definitive diagnosis of SARS-CoV-2 infection. The primary end-point was to evaluate the association of lipid markers with 30-days all-cause mortality in COVID-19.

**Results:** A total of 654 patients were enrolled, with an estimated 30-day mortality of 22.8% (149 patients). Non-survivors had lower total cholesterol (TC) and low-density lipoprotein cholesterol (LDL-c) levels during the entire course of the disease with complete resolution among survivors. Both showed a significant inverse correlation with inflammatory markers and a positive correlation with lymphocyte count. In a multivariate analysis, LDL-c ≤ 69 mg/dl (hazard ratio [HR] 1.94; 95% confidence interval [CI] 1.14-3.31), C-reactive protein > 88 mg/dl (HR 2.44; 95% CI, 1.41-4.23) and lymphopenia < 1,000 (HR 2.68; 95% CI, 1.91-3.78) at admission were independently associated with 30-day mortality. This association was maintained 7 days after admission.

**Conclusion:** Hypolipidemia in SARS-CoV-2 infection may be secondary to an immune-inflammatory response, with complete recovery in survivors. Low LDL-c serum levels are independently associated with higher 30-day mortality in COVID-19 patients.

## 1. INTRODUCTION

Severe acute respiratory syndrome coronavirus 2 (SARS-CoV-2) is a novel single-stranded RNA virus [1], and it is considered the sole causative agent of the Coronavirus disease-2019 (COVID-19), which the World Health Organization granted with the pandemic status. Over 1,016,986 patients are currently dead after 34,161,721 confirmed cases worldwide [2]. Initial reports have suggested that SARS-CoV-2 can trigger an intense inflammatory response after dysregulation of the host immune system. SARS-CoV-2 binds to a human angiotensin-converting enzyme-2 receptor to gain intracellular entry [1] after it causes some protein and lipid conformational changes at the edges of cholesterol-rich lipid domains [3].

Cholesterol is a precursor of steroid hormones and bile acids and is an essential constituent of cell membranes that facilitates signal transduction. It is considered a key mediator for cell invasion by microbial pathogens, and the acute or chronic phases of infection can alter their levels. In particular, in the acute phase of influenza infection, high-density lipoprotein cholesterol (HDL-c) can gain some pro-inflammatory properties [4]. On the other hand, lipid levels abnormalities have been described in both human immunodeficiency virus [5] and hepatitis C virus [6] infections during their chronic phases. The mechanism of alteration involves the virus to the Scavenger receptor B type 1 that facilitates the selective uptake of HDL-c and other lipid components of receptor-bound HDL particles, including free cholesterol and triglycerides (TG) [7]. Besides, membrane cholesterol is a critical component to facilitate viral entry into host cells [8].

The role of lipid metabolism in coronavirus disease has been investigated even before the current pandemic [9]. However, until now, little is known about the relationship between cholesterol and SARS-CoV-2 viral infection or COVID-19. Herein, in this study, we explored whether SARS-CoV-2 infection interferes with lipid metabolism and its prognostic during short-term follow-up.

## 2. MATERIALS AND METHODS

### 2.1 Study design and population

We conducted this ambispective single-center study at a Spanish tertiary hospital (Hospital Clínico Universitario de Valladolid, Spain, Valladolid). Between March 1^st^, 2020, and May 15^th,^ 2020, data from a total of 654 consecutive patients with a definitive diagnosis of SARS-CoV-2 infection confirmed through positive reverse transcriptase-polymerase chain reaction (RT-PCR) were prospectively collected. Exclusion criteria were age < 18 years old, pregnant women, and death in the first 24 h after admission. We retrospectively collected the baseline clinical features, radiologic procedures, and laboratory tests. We also recorded ongoing treatment and clinical outcomes from the electronic medical records. Special attention was given to the accurate recording of the prescribed therapy before admission and during hospital stay according to our institution’s protocols and discretion of the medical team. Follow-up continued through until July 25^th^, 2020. The local ethics committees approved the study protocol. Informed consent was waived, given the ambispective and observational nature of the study. The work was carried out by following the guidelines of the Declaration of Helsinki of the World Medical Association.

### 2.2 Outcome measure

The primary endpoint was to investigate any relationship between serum levels of lipid markers in the first hours of admission and all-cause mortality within 30-days. Secondary endpoints were to determine (1) the correlation between lipid markers at admission with other inflammatory markers and (2) to monitor the changes in lipid mediator’s levels during hospitalization as well as their correlation with the prognosis.

Following the institution’s protocols, a blood sample with a metabolic and lipid profile was drawn on the first 24 hours and the 7^th^ day during admission according to medical criteria. We assessed fasting total cholesterol (TC), high-density lipoprotein cholesterol (HDL-c), low-density lipoprotein cholesterol (LDL-c), and triglycerides (TG). We also recorded lipid profiles from the last eighteen months before the index event and during follow-up at the discretion of the general practitioner if available to evaluate lipid metabolism further.

### 2.3 Clinical laboratory tests

We carried out all tests at our certified clinical laboratory (ISO 9001:2015). White blood cells, lymphocyte, and monocyte counts were performed on Sysmex XN-1000® analyzer using the manufacture’s reagents (Sysmex Corporation, Japan). Alanine aminotransferase (ALT), aspartate aminotransferase (AST), gamma-glutamyl transferase, lactate dehydrogenase (LDH), LDL-c, HDL-c, TC, and TG were tested on Roche Cobas 8100 sampling system analyzer (Module Cobas® c 702, Roche Diagnostics, Switzerland) using manufacture’s reagents. The methodology for direct LDL-c, HDL-c, and TC methods are based on a standard homogeneous enzymatic colorimetric assay. We tested interleukin 6 (IL6) on IMMULITE® 2000 immunoassay system using manufacture’s luminescent immunoassay reagents (IMMULITE® 2000 IL6, Siemens Healthcare Diagnostic, Germany). Procalcitonin (PCT) measurement in plasma was performed by electrochemiluminescence immunoassay on a chemistry analyzer (Cobas 6000, Roche Diagnostics) limit of detection, as below 0.02 ng/ml. We measured serum C-reactive protein (CRP) by particle enhanced immunoturbidimetric (e501 Module Analyser, Roche Diagnostics). The limit of detection was set below 0.3 mg/L.

### 2.4 Statistical analysis

We reported categorical variables as absolute values and percentages. Continuous variables were reported as median (interquartile range [IQR]). The normal distribution of continuous variables was verified with the Kolmogorov-Smirnov test and q-q plot. Categorical variables were compared with the chi-square test and the Fisher exact test when necessary. We compared continuous variables with the Mann-Whitney U test. A Spearman test was performed to analyze the correlation between lipid variables with the rest of the serum markers. We assessed the accuracy of analyzed variables to identify non-survivors patients by using the area under the receiver operating characteristic curve analysis (AUROC). We determined the optimal operating point (OOP) in the AUROC as the one that equaled sensitivity and specificity regarding mortality, and we used it as the cut-off point in the lipid profiles. We analyzed time to 30-day mortality by Kaplan–Meier survival curves and compared using the log-rank test. In a further step, we performed a multivariable Cox-regression analysis to determine the predictors of 30-day mortality in the global study population. Proportional hazard assumptions were verified by Shoenfeld residual test and check with using the log(-log(survival)) plots. We performed the statistical analyses with the use of R software, version 3.6.1 (R Project for Statistical Computing) and IBM SPSS Statistics, Version 25.0. Armonk, NY: IBM Corp. Differences were statistically significant when the p-value was < 0.05.

## 3. RESULTS

### 3.1 Baseline and clinical characteristics

Main baseline and clinical characteristics at admission are summarized in ***Table 1***. A total of 654 patients were admitted due to COVID-19 with an estimated 30-day fatality rate of 22.8%. Non-survivors were older (82 vs. 66 years old; p<0.001), had lower baseline oxygen saturation levels at admission (91 vs 95; p<0.001), developed a significant higher degree of respiratory failure (92.5% vs. 43.3%; p<0.001) and required ICU admission more frequently (20.3% vs. 8.1%; p<0.001) than survivors. Non-survivors showed a greater prevalence of hypertension (74.5% vs. 46.7%; p<0.001), diabetes mellitus (31.5% vs 17%; p<0.001), dyslypemia (52.3% vs. 34.3%; p<0.001) as well as other comorbidities such as chronic kidney disease (19.5% vs. 5.7%; p<0.001) or ischemic heart disease (14.8% vs. 8.1%; p= 0.016). In accordance, non-survivors were more commonly treated prior to admission with antihypertensive drugs, oral antidiabetics, antiplatelets and statins. Time from symptom onset to admission was shorter (4 vs. 7 days; p<0.001) in the non-surviving cohort. They also displayed greater levels parameters of organ damage and inflammation such as creatinine (1.16 vs. 0.81 mg/dL; p<0.001), D-dimer (1,394 vs. 664 ng/ml; p <0.001), C-reactive protein (128.4 vs. 54.5 mg/dL; p<0.001), procalcitonin (0.33 vs. 0.08 ng/mL; p< 0.001), LDH (357 vs. 265 UI/L; p<0.001) and increased IL6 levels (52.1 vs. 18.4 pg/mL; p<0.001). In contrast, non-survivors had lower blood count of haemoglobin [12.4 vs. 13.3 g/dL; p<0.001) lymphocytes [805 vs. 1,130 cells/mm^3^); p<0.001) and platelets [183 vs. 218 cells/mm^3^×10^3^; p <0.001).

**Table-1.** Baseline Characteristics and main features during admission in hospitalized patients due to Coronavirus Disease 2019 according to mortality.

| Variable | All population<br>N= 654 | Survivors<br>N= 505 (77.2%) | Non-survivors<br>N= 149 (22.8%) | p-value |
| --- | --- | --- | --- | --- |
| **Demographics** |  |  |  |  |
| Female sex (%) | 278 (42.5%) | 223 (44.6%) | 55 (36.9%) | 0.116 |
| Age (years) | 70 [58-81] | 66 [55-76] | 82 [72-87] | **<0.001** |
| Cancer (%) | 117 (17.9%) | 81 (16%) | 36 (24.2%) | **0.023** |
| CKD (%) * | 58 (8.9%) | 29 (5.7%) | 29 (19.5%) | **<0.001** |
| Diabetes (%) | 133 (20.3%) | 86 (17%) | 47 (31.5%) | **<0.001** |
| Dyslipemia (%) | 251 (38.4%) | 173 (34.3%) | 78 (52.3%) | **<0.001** |
| Hypertension (%) | 347 (53.1%) | 236 (46.7%) | 111 (74.5%) | **<0.001** |
| Hypothyroidism (%) | 81 (12.4%) | 51 (10.1%) | 22 (13.4%) | 0.252 |
| IHD (%) | 63 (9.6%) | 41 (8.1%) | 22 (14.8%) | **0.016** |
| Obesity (%) | 62 (9.5%) | 47 (9.3%) | 15 (10.1%) | 0.781 |
| Prior pulmonary disease (%) | 143 (21.9%) | 106 (21%) | 37 (24.8%) | 0.319 |
| Prior rheumatic disease (%) | 48 (7.3%) | 34 (6.7%) | 14 (9.4%) | 0.273 |
| Prior stroke/TIA (%) | 41 (6.3%) | 29 (5.9%) | 12 (8.1%) | 0.306 |
| **Treatment prior to admission** |  |  |  |  |
| Antiplatelets (%) | 106 (16.2%) | 70 (13.9%) | 36 (24.3%) | **0.002** |
| ACEI/ARB (%) | 266 (40.7%) | 182 (36%) | 84 (56.8%) | **<0.001** |
| Levothyroxine (%) | 71 (10.9%) | 52 (10.3%) | 19 (12.8%) | 0.383 |
| Oral anticoagulation (%) | 79 (12.1%) | 40 (8.3%) | 37 (25%) | **<0.001** |
| Oral antidiabetics (%) | 106 (16.2%) | 73 (14.5%) | 33 (22.3%) | **0.023** |
| Statins (%) | 210 (32.2%) | 151 (29.9%) | 59 (39.9%) | **0.022** |
| **Main findings at admission** |  |  |  |  |
| Time from onset (days) | 7 [3-10] | 7 [4-10] | 4 [2-7] | **<0.001** |
| Pathological Chest X-ray (%) | 597 (91.8%) | 462 (91.8%) | 135 (91.8%) | 0.996 |
| Positive RT-PCR (%) ** | 638 (97.6%) | 489 (96.8%) | 149 (100%) | **0.030** |
| Basal O<sup>2</sup> saturation (%) | 94 [91-96] | 95 [92-97] | 91 [86-94] | **<0.001** |
| Sat:FiO<sup>2</sup> | 447 [433-457] | 452 [438-462] | 433 [409-448] | **<0.001** |
| **Laboratory findings at admission** |  |  |  |  |
| Haemoglobin (g/dL) | 13.1 [12-14.4] | 13.3 [12.2-14.5] | 12.4 [11-14] | **<0.001** |
| CK (mg/dL) | 80 [49-134] | 78 [49-123] | 101 [53-221] | **0.016** |
| C-Reactive protein (mg/L) | 67.7 [26.75-134.05] | 54.5 [21-105.6] | 128.4 [72.2-191.5] | **<0.001** |
| Creatinine (mg/dL) | 0.87 [0.7-1.15] | 0.81 [0.7-1.02] | 1.16 [0.88-1.63] | **<0.001** |
| D-Dimer (ng/mL) | 756 [449-1,386] | 664 [414-1,134] | 1,394 [747-2,576] | **<0.001** |
| Ferritin (ng/mL) | 609 [294-1,134] | 605 [290-1,100] | 640 [302-1,298] | 0.205 |
| ALT (U/L) | 24 [17-43] | 25 [17-44] | 22 [15-37] | **0.004** |
| AST (UI/L) | 36 [24-52] | 34 [24-49] | 41 [25-60] | **0.022** |
| Interleukin-6 (pg/mL) | 20.9 [10-47] | 18.4 [9.3-40.5] | 52.1 [22.5-115] | **<0.001** |
| LDH (UI/L) | 283 [215-359] | 265 [206-331] | 357 [303-460] | **<0.001** |
| Lymphocytes (cells/mm<sup>3</sup>) | 1,050 [720-1,470] | 1,130 [820-1,570] | 805 [580-1,215] | **<0.001** |
| Neutrophils (cells/mm<sup>3</sup>x10<sup>3</sup>) | 4,600 [3,020-6,830] | 4,260 [2,840-5,920] | 6,125 [4,035-9,805] | **<0.001** |
| Platelets (cells/mm<sup>3</sup>x10<sup>3</sup>) | 213 [163-274] | 218 [169-284] | 183 [137-260] | **<0.001** |
| Procalcitonin (ng/ml) | 0.11 [0.06-0.29] | 0.08 [0.05-0.16] | 0.33 [0.13-0.82] | **<0.001** |
| Troponin (ng/L) | 15.04 [7.91-22.28] | 9.3 [7-17.2] | 33.6 [16-148.1] | **<0.001** |
| **Specific COVID-19 treatment** |  |  |  |  |
| **Azithromycin (%)** | 588 (94.2%) | 470 (95.3%) | 118 (90.1%) | **0.022** |
| **Betaferon (%)** | 244 (39.1%) | 182 (36.9%) | 62 (47.3%) | **0.03** |
| **Hydroxychloroquine (%)** | 605 (97%) | 483 (98%) | 122 (93.1%) | **0.008** |
| **Lopinavir/Ritonavir (%)** | 590 (94.4%) | 473 (95.7%) | 117 (89.3%) | **0.004** |
| **Non-specific COVID-19 treatment** |  |  |  |  |
| **Antibiotics (%)** | 551 (84.4%) | 424 (84.1%) | 127 (85.2%) | 0.743 |
| **Anticoagulation (%) †** | 545 (83.8%) | 434 (86.3%) | 111 (75.5%) | **0.002** |
| **Corticosteroids/Glucocorticoids (%)** | 383 (58.6%) | 289 (57.2%) | 94 (63.1%) | 0.202 |
| **Statins (%)** | 48 (7.4%) | 37 (7.4%) | 11 (7.4%) | 0.996 |
| **Main in-hospital outcomes** |  |  |  |  |
| **LOS (days)** | 9 [5-15] | 10 [6-16] | 7 [4-14] | **<0.001** |
| **ICU admission (%)** | 71 (10.9%) | 41 (8.1%) | 30 (20.3%) | **<0.001** |
| **Mechanical ventilation (%)** | 77 (9.7%) | 33 (6.6%) | 34 (22.1%) | **<0.001** |
| **Nosocomial infection (%)** | 122 (18.7%) | 75 (14.9%) | 47 (31.5%) | **<0.001** |
| **Respiratory failure (%) ††** | 354 (54.5%) | 218 (43.3%) | 136 (92.5%) | **<0.001** |

ACEi: Angiotensin-converting enzyme inhibitors; ALT: alanine aminotransferase; ARB: angiotensin receptor blocker; AST: aspartate aminotransferase CKD: Chronic kidney disease; ICU: Intensive care unit; IHD: Ischemic heart disease; LDH: Lactate dehydrogenase; LOS: Length of stay; RT-PCR: Reverse transcription-polymerase chain reaction;
\* Chronic kidney disease was defined as a glomerular filtration rate of <60 ml/min or need for dialysis \*\* At hospital admission; † includes prophylactic, intermediate and complete anticoagulation doses; †† Defined as pO<sub>2</sub> <60 mmHg, SO<sub>2</sub> <92% or need for non-invasive or mechanical ventilation

Specific COVID-19 treatment was more commonly prescribed in survivors with comparable prescription rate in respect to statins (7.4% vs. 7.4%; p= 0.996) and corticosteroids (57.2% vs. 63.1%; p= 0.202). Nosocomial infection (31.5% vs. 14.9%; p<0.001) were more common among non-survivors during hospitalization. ***Table 1*** shows the main clinical outcomes and laboratory findings during hospitalization.

### 3.2 Lipid profile among COVID-19 patients

Lipid profiles were tracked from previous laboratory reports before SARS-CoV-2 infection (when available), at hospital admission, at 7^th^ day during hospitalization, and until death or first-time follow-up after discharge (***see Figure 1 and Table 2***). In the overall population (including survivors and non-survivors), serological levels of all the lipid markers analyzed, except TG, displayed a significant decrease at the time of admission concerning the previous baseline values. Besides, baseline serum TC and LDL-c levels were significantly higher in survivors than non-survivors at any time; whereas, HDL-c was comparable at admission but significantly lower in non-survivor before (47.2 vs. 52.6 ng/dL; p = 0.004) and in the 7^th^ day (27 vs. 34 mg/dL; p= 0.011) after hospital admission. Survivors had a progressive recovery of TC, LDL-c, and HDL-c levels; however, this was not the case in the non-survivor cohort. Finally, median follow-up among survivors was 73 days with complete recovery to previous basal levels of their lipid profiles (***see Figure 1 and Table 2***).

**Table-2.** Lipid profile of patients with Coronavirus Disease 2019 in the global study population and according to mortality during the full course of the disease.

| Variable | All population<br>N= 654 | Survivors<br>N= 505 (77.2 %) | Non-survivors<br>N= 149 (22.8%) | p-value |
| --- | --- | --- | --- | --- |
| **Lipid laboratory profile before admission** |  |  |  |  |
| Total cholesterol (mg/dL) | 187.5 [155-214] | 191 [164.5-215] | 167.5 [133-200] | **<0.001** |
| HDL-c (mg/dL) | 51.6 [41.7-62.1] | 52.6 [43-64] | 47.2 [37.5-60] | **0.004** |
| LDL-c (mg/dL) | 108.4 [84.6-129.6] | 113.2 [91.3-134.3] | 93 [66.4-112.6] | **<0.001** |
| Triglycerides (mg/dL) | 109.5 [82.0-146.0] | 108 [79-146] | 116.5 [91-145] | 0.122 |
| TC/HDL-c | 3.5 [2.9-4.3] | 3.5 [3-4.3] | 3.5 [2.8-4] | 0.328 |
| **Lipid laboratory profile at admission** |  |  |  |  |
| Total cholesterol (mg/dL) | 137 [117-163] | 142 [120-165] | 121 [101.5-146.5] | **<0.001** |
| HDL-c (mg/dL) | 34 [28-43] | 34 [28.3-43] | 33.5 [27-44] | 0.278 |
| LDL-c (mg/dL) | 77 [59-97] | 80 [63-98] | 61.1 [41-84.2] | **<0.001** |
| Triglycerides (mg/dL) | 118 [91-163] | 119 [92-163] | 114 [86.5-157] | 0.240 |
| TC/HDL-c | 3.9 [3.11-4.84] | 3.9 [3.2-4.9] | 3.39 [2.9-4.6] | **0.012** |
| **Lipid laboratory profile during 7<sup>th</sup> day of hospitalization** |  |  |  |  |
| Total cholesterol (mg/dL) | 157 [126-192] | 163 [130.5-197] | 134 [106.5-158] | **<0.001** |
| HDL-c (mg/dL) | 33 [26.5-41.6] | 34 [28-41.8] | 27 [19.1-37] | **0.011** |
| LDL-c (mg/dL) | 79.7 [55-107.5] | 85.7 [62-115.5] | 56.4 [39.3-75.1] | **0.001** |
| Triglycerides (mg/dL) | 184 [130-257] | 189 [132-261.5] | 161.5 [124-226] | 0.057 |
| TC/HDL-c | 4.54 [3.73-5.95] | 4.5 [3.7-5.9] | 4.4 [3.6-6.1] | 0.965 |
| **Lipid laboratory profile during follow-up \*** |  |  |  |  |
| Total cholesterol (mg/dL) | 196 [168-223.5] | 196 [168-223.5] | NA | NA |
| HDL-c (mg/dL) | 52.8 [44.1-64] | 52.8 [44.1-64] | NA | NA |
| LDL-c (mg/dL) | 117.3 [99.3-144.7] | 117.3 [99.3-144.7] | NA | NA |
| Triglycerides (mg/dL) | 111 [83-156] | 111 [83-156] | NA | NA |
| TC/HDL-c | 3.7 [3.2-4.3] | 3.7 [3.2-4.3] | NA | NA |

HDL High-density lipoprotein cholesterol; LDL-c: Low-density lipoprotein cholesterol; NA: Not applicable; TC: Total cholesterol; TG Triglycerides
\* Only in survivors

**Figure 1.**
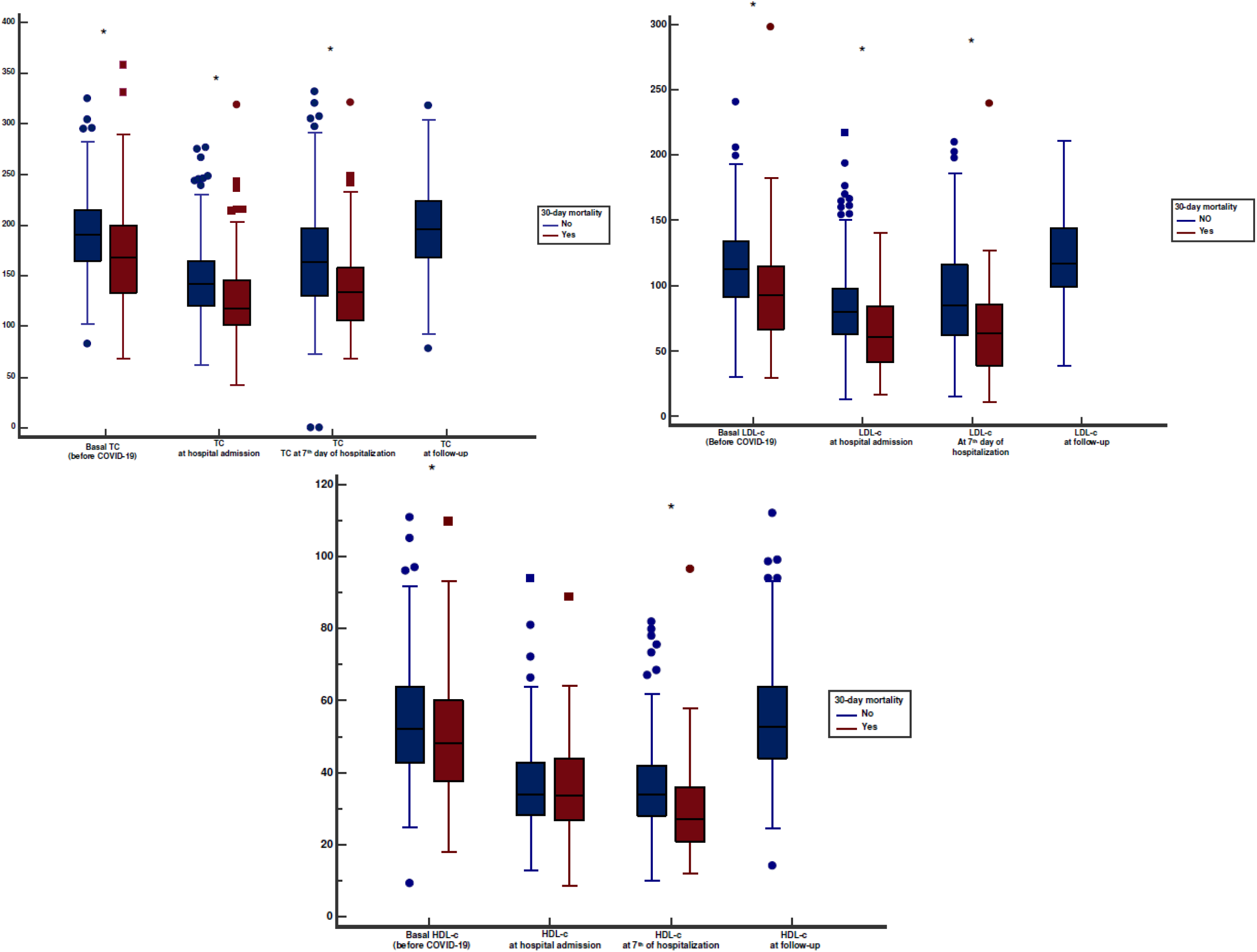
Temporal changes in lipid profile levels in COVID-19 patients according to the clinical course of the disease: **A)** Total cholesterol; **B)** LDL-c; **C)** HDL-c. The horizontal lines represent the median value in each group. HDL-c: High-density lipoprotein cholesterol; LDL-c: Low-density lipoprotein cholesterol; TC: Total cholesterol **\*** p <0.05

**Figure 2.**
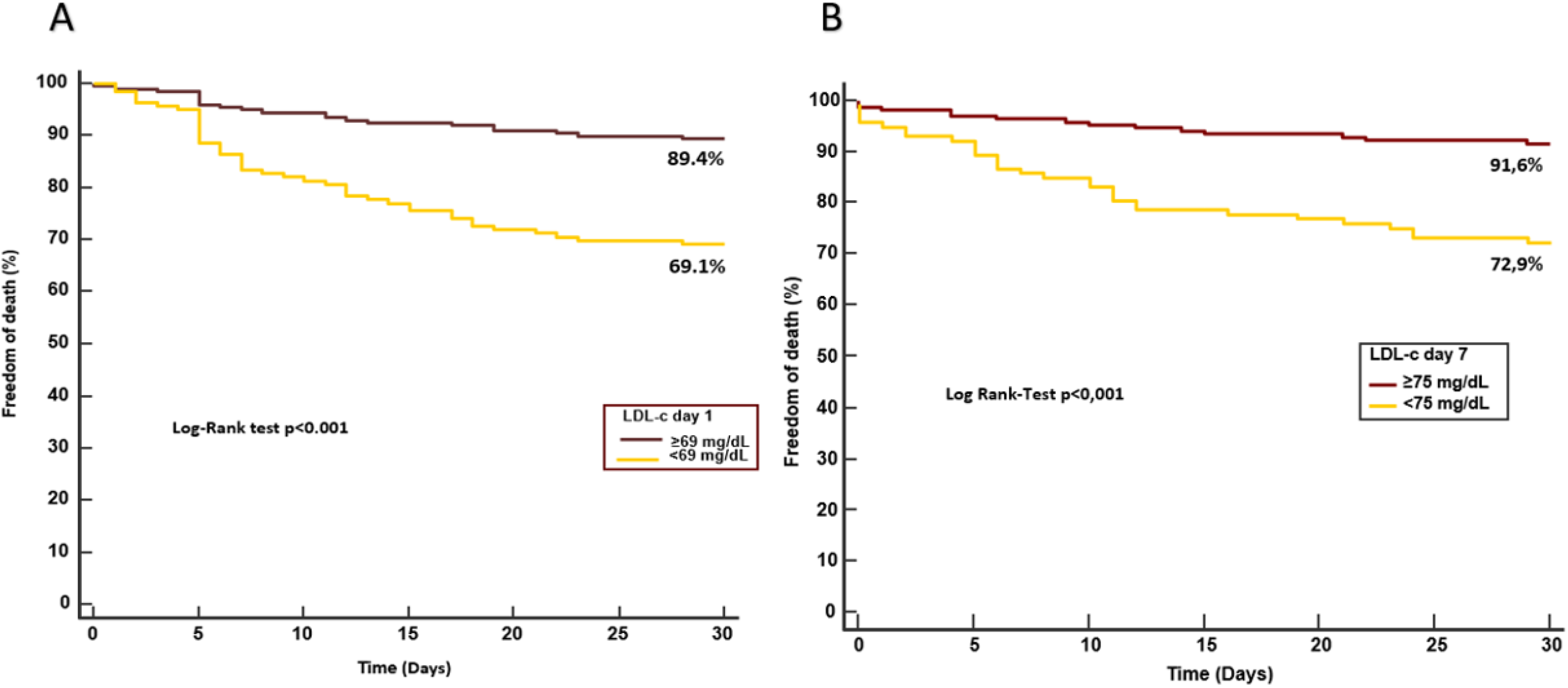
Kaplan-Meier estimates of mortality in the total COVID-19 population according to LDL-c **A)** at admission and **B)** at the 7^th^ day of hospitalization based on their optimal cut-off point.

### 3.3 Correlation of lipid markers with other inflammatory markers

A Spearman correlation analysis assessed the relationship of lipid parameters with all the gathered analytical parameters. Interestingly, LDL-c and TC levels at admission were inversely correlated with the levels of CRP (r= −0.217 and −0.209, respectively; p<0.001), PCT (r= −0.313 and − 0.229; p <0.001) and IL6 (r=-0.334 and −0.301; p<0.001) with a positive correlation with lymphocytes (r=0.179 and 0.191; p= 0.001), which was maintained throughout hospitalization and until recovery (***see supplementary Table 1***). LDL-c did show a very strong positive correlation with TC (r= 0.937; p<0.001) as opposed to HDL-c (r= 0.201; p<0.001).

### 3.4 AUROC curve analysis

We analyzed the diagnostic performance accuracy of lipid profiles to predict 30-day mortality (***see supplementary Figure 1***). Only LDL-c and TC showed a significant AUROC during admission and 7^th^ day of hospitalization; in particular, LDL-c showcased better AUROC than TC regardless time course of the disease. Hence, AUROC curve for LDL-c at admission was 0.7 (95% CI 0.64-0.75; p<0.001) as opposed to TC with an AUROC of 0.67 (95% CI 0.63-0.71; p<0.001). On the 7^th^ day, AUROC for LDL-c and TC were 0,75 (95% CI 0.63-0.82; p<0.001) and 0.7 (95% CI 0.6-0.77; p<0,001), respectively. The estimated threshold values for LDL-c were 69 mg/dl on admission and 75 mg/dl in the 7^th^ day of hospitalization. On the contrary, TC estimated cut-off values were 132 mg/dL and 147 mg/dL, respectively.

### 3.5 Association between mortality and lipid profile

Variables associated with 30-day mortality in multivariate analysis are summarized in ***Table 3***. Independent predictors of mortality were estimated through a Cox multivariate regression analysis. The variables included in the model were those with a p-value <0.05 on the univariate analysis (age, hypertension, diabetes, dyslypemia, ischemic heart disease, chronic renal disease, angiotensin receptor antagonist, angiotensin converting enzyme inhibitors, statins, lymphopenia, c-reactive protein, antiviral treatment, anticoagulation, total cholesterol and LDL-c). It revealed that age (hazard ratio [HR] 1.08; 95% confidence interval [CI], 1.05-1.11]; p <0.001), lymphopenia <1,000 cells/ml (HR 2.68; 95% CI, 1.91-3.78; p<0.001), LDL-c <69 mg/dL (HR 1.94; 95% CI, 1.14-3.31; p = 0.014) and CRP >88 mg/dL (HR 2.44; 95% CI, 1.41-4.23; p= 0.001) at admission were independent variables associated with a greater risk of 30-day mortality. Those findings remained consistent on the seventh day of hospitalization with a cut-off of 75 mg/dL and 33 mg/dL for LDL-c and CRP, respectively. Unadjusted survival Kaplan-Meier curves for 30-day global mortality was performed and shown in ***Figure 3***, those patients with LDL-c levels <69 mg/dl at the time of admission and <75 mg/dl on the 7^th^ day showed a 20% higher 30-day mortality rate than the rest of the patients.

**Table-3.**
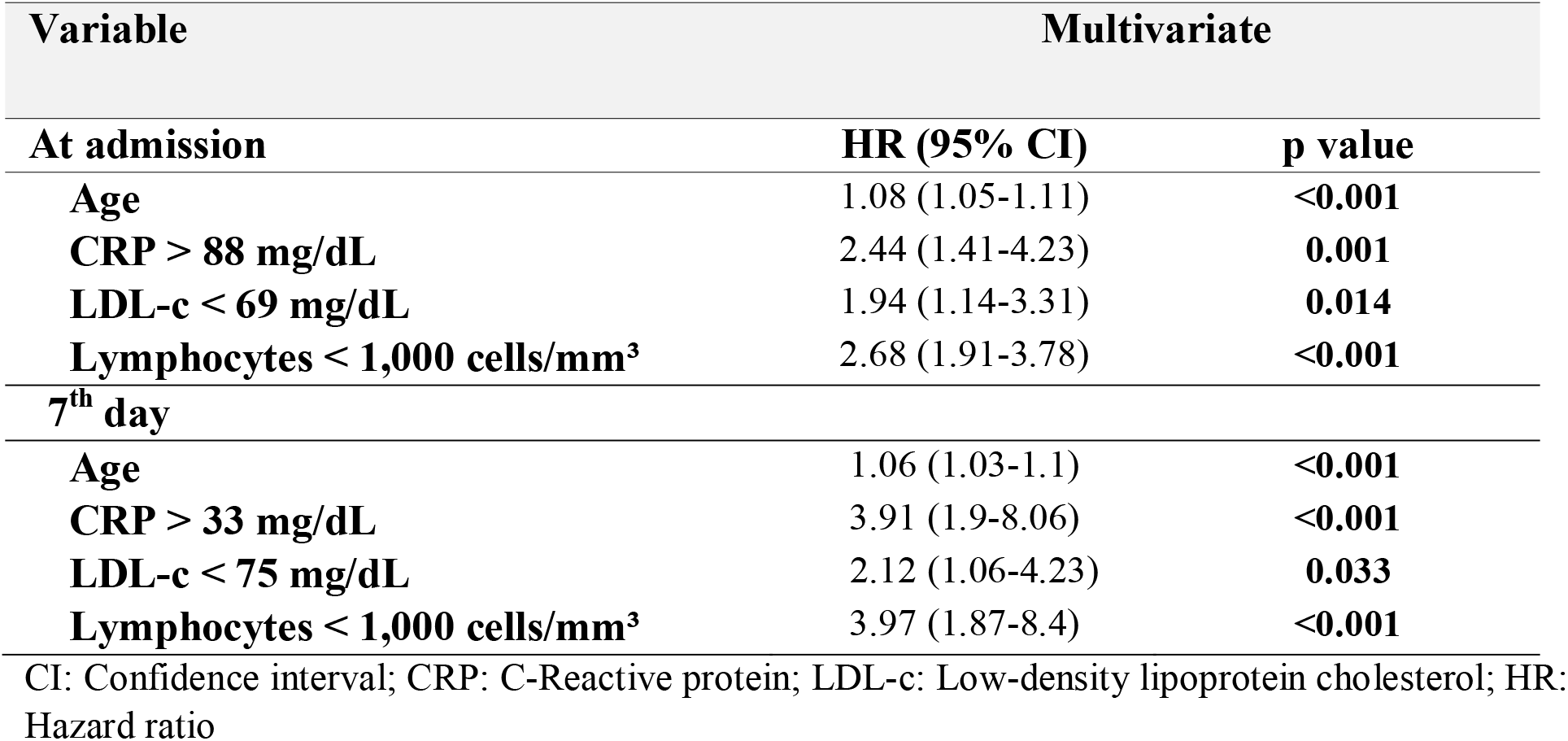
Multivariate Cox regression analysis for evaluating the risk of 30-day mortality in the COVID- 19 study population.

## 4. DISCUSSION

Lipoproteins play a vital role in the innate immunity and perform different functions against infection: receptor blocking, lysis, chemotaxis, and neutralization of bacterial endotoxins. Lipids are essential for the replication and pathogenicity of enveloped viruses [10,11]. The main findings of this research are: (1) the presence of immune-inflammatory mediated hypolipidemia in SARS-CoV-2 infection; and (2) the role of LDL-c as a potential prognostic marker in septic patients, COVID-19 patients with low LDL-c were more likely to die.

The mechanism behind the observed hypolipidemia is likely multifactorial and complex. Serum levels of ALT, AST, GGT, and LDH increased moderately in non-surviving patients, indicating mild liver-function damage, which might be a contributing factor to hypolipidemia by disrupting uptake and biosynthesis of lipoproteins [12]. Nonetheless, a specific type of viral infections can lead to alteration of lipid metabolism in the acute and chronic phases as a response of an ongoing inflammatory state [4,5].

Although COVID-19 pathophysiology is not fully understood, COVID-19 severity and death are associated with a hyperinflammatory state due to a dysregulated immune system [13]. The clinical profile of the patients included in this study shows a similar trend with systemic inflammation being a notable contributor to mortality, but also of SARS-CoV-2 mediated dyslipidemia. We have observed a paradoxical lipid metabolism with a U-shaped curve of lipid levels with similar findings previously described in inflammatory diseases [14]. Several are the plausible mechanism from an immunological point of view.

HDL-c might become pro-inflammatory under specific conditions that increase reactive oxygen species and myeloperoxidase activity. It can also modify their levels and Apoprotein-AI concentration; hence, altered reverse cholesterol transport [4,15]. LDL-c can be oxidized when its HDL-c counterpart loses its antioxidative properties, or if oxidized phospholipids accumulate. They are identified as damaged-associated molecular patters (DAMPs) by scavenger receptors, activate the inflammasome [4] and the immune system [13]. Low LDL-c levels may also be the consequence of an increase in vascular leakage into the lung parenchyma as a result of endothelial damage [12,13].

Finally, an increased concentration of pro-inflammatory cytokines may be responsible for a drastic decrease in plasma LDL-c levels during the acute-phase response. Direct effects of cytokines might explain the altered lipid concentrations [14] by up-regulating ox-LDL uptake or overriding suppression of LDL-c receptor through the expression of scavenger receptors. These changes observed with inflammation can increase the odds of cardiovascular disease through the formation of foam cells and endothelial damage [15, 16].

Sepsis is defined as the presence of infection with a detrimental host response with organ damage in the most critical cases [17]. Low HDL-c levels have been associated before with an increased risk of sepsis [18,19] and adverse outcomes [20-23]. In fact, Maile et al. [24] or Guirgis et al. [25] also suggested that low baseline LDL-c levels are associated with an increased risk of mortality and sepsis, respectively. By analogy, similar findings should be identified in SARS-CoV-2 patients.

In particular, low HDL-c levels in SARS-CoV-2 patients have been associated with disease severity [26, 27], but our results are in agreement with those of recently published where low LDL-c levels were associated with COVID-19 severity [12,28]. However, we also observed an association with an increased risk of mortality with low LDL-c levels after the adjusted multivariate analyses. By contrast, a recent work by Walley et al. proposed that low LDL-c levels are merely an indicator of the disease severity in septic patients without a contributing role in mortality [29].

The observed associated mortality in this cohort of COVID-19 patients may be explained by other mechanism. In this sense, LDL-c carries a large percentage of plasma Coenzyme Q10 (CoQ10) that has significant antioxidant capacity, avoiding peroxidative damage to the cellular membranes [30, 31]. Low LDL-c levels can cause a decrease in plasma CoQ10 levels, which can lead to endothelial dysfunction, organ damage, and death as observed in COVID-19 patients [32]. All these mechanisms justify that patients with low LDL-c levels have a reduced defensive, energetic and metabolic reaction capacities to be able to properly manage a situation of aggression and organ stress such as COVID-19, thus associating a significant increase in mortality.

Overall, low LDL-c levels may reflect the acute phase of the disease, but they may also induce multiple systemic reactions through a complex interplay. Therefore, in the appropriate scenario, we might hypothetically consider low LDL-c levels as a plausible candidate as a routine risk marker during admission and also in the first weeks of disease evolution. Nevertheless, we cannot rule out a catabolic state or high immune cell turnover as a cause of hypolipidemia in COVID-19 patients. The presence in our study of a statistically significant positive correlation between LDL-c levels and lymphocyte count in blood, the latter being an independent variable associated with 30-day mortality together with LDL-c in multivariate regression analysis, can support this theory.

Our work presents certain limitations. These observations should be considered hypothesis-generating only due to the intrinsic retrospective nature of the present work. Moreover, the data were subject to selection bias, and the generalizability of the results may be reduced by the fact that we did not evaluate outpatients. We could not measure apoproteins or oxidized forms of main lipoproteins, which may play a detrimental role in the pathogenesis of COVID-19. Finally, for better characterization of this hypolipidemia, our findings should be validated in a large multicentric cohort of COVID-19 patients monitoring the dynamics of lipid profiles before, during the acute phase and follow-up.

In conclusion, our results suggest that COVID-19 hypolipidemia is associated with inflammation; In particular, LDL-c could be used as a complementary marker in septic patients for better risk stratification. Upcoming studies should determine to what extent treatment-related resolution of inflammation or changes in lipid levels may impact outcomes; such data are currently lacking.

## Data Availability

All labor data in this work are real and are available in electronic format to external reviewers and external evaluators however there is no linked URL.

## ABBREVIATIONS

ALT: Alanine aminotransferase
AST: Aspartate aminotransferase
AUROC: Area under the receiver operating characteristic curve analysis
CoQ10: Coenzyme Q10
COVID-19: Coronavirus disease 2019
CRP: C-Reactive protein
HDL-c: High-density lipoprotein cholesterol
IL6: Interleukin 6
LDH: Lactate dehydrogenase
LDL-c: Low-density lipoprotein cholesterol
PCT: Procalcitonin
RT-PCR: Reverse transcription-polymerase chain reaction
SARS-CoV-2: Severe acute respiratory syndrome coronavirus 2
TC: Total cholesterol
TG: Triglycerides

## REFERENCES

1. Madjid M, Safavi-Naeini P, Solomon SD, Vardeny O. Potential Effects of Coronaviruses on the Cardiovascular System. Jama Cardiol. 2020; 5(7).

2. WHO. Coronavirus disease 2019 (COVID-19) situation report. October 3, 2020. https://www.who.int/emergencies/diseases/novel-coronavirus-2019 (accessed October 3, 2020).

3. Radenkovic D, Chawla S, Pirro M, Sahebkar A, Banach M. Cholesterol in Relation to COVID-19: Should We Care about It? J Clin Medicine. 2020; 9(6):1909.

4. Tall AR, Yvan-Charvet L. Cholesterol, inflammation and innate immunity. Nat Rev Immunol. 2015; 15(2):104–116.

5. Funderburg NT, Mehta NN. Lipid Abnormalities and Inflammation in HIV Inflection. Curr Hiv-aids Rep. 2016; 13(4):218–225.

6. Tien PC. Hepatitis C Virus-Associated Alterations in Lipid and Lipoprotein Levels: Helpful or Harmful to the Heart? Clin Infect Dis. 2017; 65(4):566–567.

7. Wei C, Wan L, Zhang Y, et al. Cholesterol Metabolism--Impact for SARS-CoV-2 Infection Prognosis, Entry, and Antiviral Therapies. Medrxiv [Internet]. 2020; :2020.04.16.20068528. Available from: http://medrxiv.org/content/early/2020/04/24/2020.04.16.20068528.abstract

8. Meher G, Bhattacharjya S, Chakraborty H. Membrane Cholesterol Modulates Oligomeric Status and Peptide-Membrane Interaction of Severe Acute Respiratory Syndrome Coronavirus Fusion Peptide. J Phys Chem B. 2019; 123(50):10654–10662.

9. Wu Q, Zhou L, Sun X, et al. Altered Lipid Metabolism in Recovered SARS Patients Twelve Years after Infection. Sci Rep-uk. 2017; 7(1):9110.

10. Yan B, Chu H, Yang D, et al. Characterization of the Lipidomic Profile of Human Coronavirus-Infected Cells: Implications for Lipid Metabolism Remodeling upon Coronavirus Replication. Viruses. 2019; 11(1):73.

11. Alsaadi EAJ, Jones IM. Membrane binding proteins of coronaviruses. Future Virol. 2019; 14(4):275–286.

12. Wei X, Zeng W, Su J, et al. Hypolipidemia is associated with the severity of COVID-19. J Clin Lipidol. 2020; 14(3):297–304.

13. Merad M, Martin JC. Pathological inflammation in patients with COVID-19: a key role for monocytes and macrophages. Nat Rev Immunol. 2020; 20(6):355–362.

14. Robertson J, Peters MJ, McInnes IB, Sattar N. Changes in lipid levels with inflammation and therapy in RA: a maturing paradigm. Nat Rev Rheumatol. 2013; 9(9):513–523.

15. Rosenson RS, Brewer HB, Ansell BJ, et al. Dysfunctional HDL and atherosclerotic cardiovascular disease. Nat Rev Cardiol. 2016; 13(1):48–60.

16. Esteve E, Ricart W, Fernández-Real JM. Dyslipidemia and inflammation: an evolutionary conserved mechanism. Clin Nutr. 2005; 24(1):16–31.

17. Singer M, Deutschman CS, Seymour CW, et al. The Third International Consensus Definitions for Sepsis and Septic Shock (Sepsis-3). Jama. 2016; 315(8):801–810.

18. Madsen CM, Varbo A, Tybjærg-Hansen A, Frikke-Schmidt R, Nordestgaard BG. U-shaped relationship of HDL and risk of infectious disease: two prospective population-based cohort studies. Eur Heart J. 2017; 39(14):1181–1190.

19. Feng Q, Wei W-Q, Chaugai S, et al. Association Between Low-Density Lipoprotein Cholesterol Levels and Risk for Sepsis Among Patients Admitted to the Hospital With Infection. Jama Netw Open. 2019; 2(1):e187223.

20. Kaysen GA, Ye X, Raimann JG, et al. Lipid levels are inversely associated with infectious and all-cause mortality: international MONDO study results. J Lipid Res. 2018; 59(8):1519–1528.

21. Tanaka S, Couret D, Tran-Dinh A, et al. High-density lipoproteins during sepsis: from bench to bedside. Crit Care. 2020; 24(1):134.

22. Tanaka S, Diallo D, Delbosc S, et al. High-density lipoprotein (HDL) particle size and concentration changes in septic shock patients. Ann Intensive Care. 2019; 9(1):68.

23. Cirstea M, Walley KR, Russell JA, Brunham LR, Genga KR, Boyd JH. Decreased high-density lipoprotein cholesterol level is an early prognostic marker for organ dysfunction and death in patients with suspected sepsis. J Crit Care. 2017; 38:289–294.

24. Maile MD, Sigakis MJ, Stringer KA, Jewell ES, Engoren MC. Impact of the pre-illness lipid profile on sepsis mortality. J Crit Care. 2020; 57:197–202.

25. Guirgis FW, Donnelly JP, Dodani S, et al. Cholesterol levels and long-term rates of community-acquired sepsis. Crit Care. 2016; 20(1):408.

26. Hu X, Chen D, Wu L, He G, Ye W. Declined serum high density lipoprotein cholesterol is associated with the severity of COVID-19 infection. Clin Chim Acta. 2020; 510:105–110.

27. Hu X, Chen D, Wu L, He G, Ye W. Low Serum Cholesterol Level Among Patients with COVID-19 Infection in Wenzhou, China. Ssrn Electron J. 2020.

28. Fan J, Wang H, Ye G, et al. Low-density lipoprotein is a potential predictor of poor prognosis in patients with coronavirus disease 2019. Metabolis. 2020; 107:154243.

29. Walley KR, Boyd JH, Kong HJ, Russell JA. Low Low-Density Lipoprotein Levels Are Associated With, But Do Not Causally Contribute to, Increased Mortality in Sepsis*. Crit Care Med. 2019; 47(3):463–466.

30. Pacanowski MA, Frye RF, Enogieru O, et al. Plasma Coenzyme Q10 Predicts Lipid-lowering Response to High-Dose Atorvastatin. J Clin Lipidol. 2008; 2(4):289–97.

31. Dupic L, Huet O, Duranteau J. Coenzyme Q10 deficiency in septic shock patients. Crit Care. 2011;15(5):194.

32. Pons S, Fodil S, Azoulay E, Zafrani L. The vascular endothelium: the cornerstone of organ dysfunction in severe SARS-CoV-2 infection. Crit Care. 2020; 24(1):353.

