## supplementary table 1 and supplementary figure 1 for "Low-density lipoprotein cholesterol levels are associated with poor clinical outcomes in COVID-19"

**SUPPLEMENTARY MATERIAL**

**Supplementary Table-1.** Correlations of circulating lipid levels with inflammatory serum markers and lymphocytes. A Spearman correlation analysis was used.

| **Variable**† | **Lymphocytes** | **C-Reactive protein** | **Ferritin** | **Interleukin-6** | **Procalcitonin** |
| --- | --- | --- | --- | --- | --- |
| **Lipid laboratory profile at admission** |  |  |  |  |  |
| **Total cholesterol (mg/dL)** | 0.191 * | -0.209 * | -0.054 | -0.301 * | -0.229* |
| **HDL-c (mg/dL)** | -0.053 | -0.228 * | -0.264 * | -0.052 | -0.189 * |
| **LDL-c (mg/dL)** | 0.179 * | -0.217 * | -0.054 | -0.334 * | -0.313* |
| **Triglycerides (mg/dL)** | 0.059 | 0.11 * | 0.257 * | -0.037 | 0.118 * |
| **Lipid laboratory profile on the 7^th^ day** | | | | | |
| **Total cholesterol (mg/dL)** | 0.203 * | -0.396 * | 0.071 | -0.266 * | -0.594* |
| **HDL-c (mg/dL)** | -0.06 | -0.315 * | -0.163 * | -0.211 | 0.361 |
| **LDL-c (mg/dL)** | 0.165 * | -0.388 * | 0.062 | 0.224 | -0.547* |
| **Triglycerides (mg/dL)** | 0.178 * | -0.132 * | 0.186 * | 0.077 | 0.210 |
| **Lipid laboratory profile during follow-up**†† | | | | | |
| **Total cholesterol (mg/dL)** | NA | -0.109 | 0.102 | -0.127 | NA |
| **HDL-c (mg/dL)** | NA | -0.205 * | -0.08 | - 0.140 | NA |
| **LDL-c (mg/dL)** | NA | -0.183 * | 0.137 | 0.019 | NA |
| **Triglycerides (mg/dL)** | NA | 0.148 * | 0.028 | 0.227 | NA |

Abbreviations: HDL: High density lipoprotein cholesterol: LDL-c: Low density lipoprotein cholesterol; NA: Not applicable; TC: Total cholesterol: TG Triglycerides

* p <0.05

† Variables where compare at the specific given moment; †† Only applicable to survivors

**Supplementary Figure 1.** AUROC analysis for predicting 30-day mortality. Accuracy of the four components of the lipid profile were analyzed upon admission (**panel A**) and during the first week (**panel B**) for differentiating between survivors and non-survivors.

Only significant AUROC values are shown in the tables.

**Abbreviations:** HDL: High density lipoprotein cholesterol: LDL-c: Low density lipoprotein cholesterol; TC: Total cholesterol: TG: Triglycerides; Sens: Sensitivity; Spc: Specificity

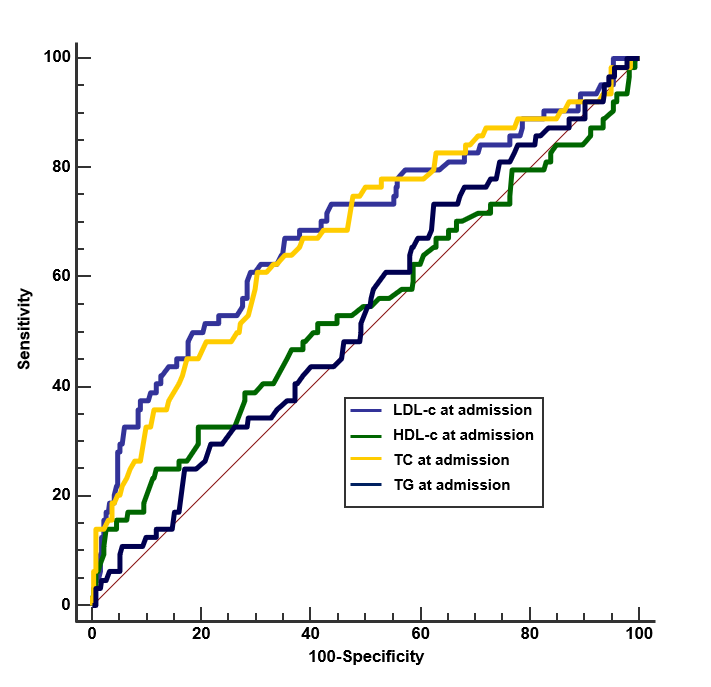

|  | **AUROC (95% IC)** | **p** | **Cut-off (mg/dl)** | **Sens** | **Spc** |
| --- | --- | --- | --- | --- | --- |
| LDL-c | 0,7 (0.64 - 0.75) | <0,001 | 69 | 0.66 | 0.68 |
| TC | 0,67 (0.63 - 0.71) | <0,001 | 132 | 0.61 | 0.68 |

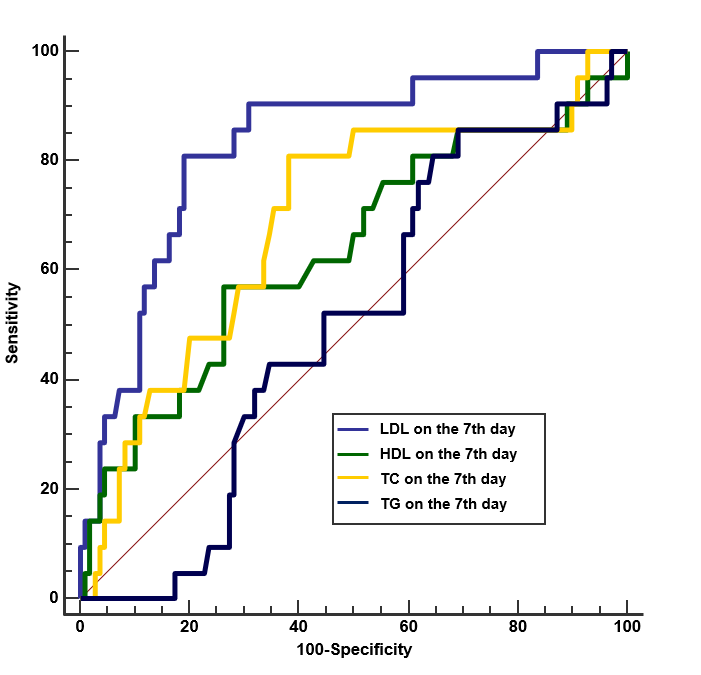

|  | **AUROC (95% IC)** | **p** | **Cutt-off (mg/dl)** | **Sens** | **Spc** |
| --- | --- | --- | --- | --- | --- |
| LDL-c | 0,75 (0.63 - 0.82) | <0,001 | 75 | 0.70 | 0.69 |
| TC | 0,70 (0.6-0.77) | <0,001 | 147 | 0.68 | 0.63 |
